## Supplementary material for "COVID-19 vaccination beliefs, attitudes, and behaviours among health and social care workers in the UK: a mixed-methods study": S1 Appendix 1

COVID-19 vaccine views, intentions and behaviours in H&SC workers

Start of Block: Block 1

Q1
 Coronavirus (COVID-19) vaccine views and experiences amongst health and social care workers in the UK
 **INVESTIGATORS AND COLLABORATORS** 
  **London School of Hygiene & Tropical Medicine**
 Sadie Bell, Pauline Paterson, & Sandra Mounier-Jack
  
**University of Southampton**
 Richard Clarke
  
**Public Health England**
 Louise Letley
  
**NHS Race and Health Observatory**
 Habib Naqvi & Yvonne Coghill
  
**Royal College of Nursing**
 Helen Donovan   **INVITATION**      You are invited to take part in an anonymous survey looking at health and social care worker views on COVID-19 vaccination. You are eligible to participate in this survey if you work in the UK in a frontline health or social care worker role (i.e. a role where you have direct contact with patients, service users, or clients). As COVID-19 has hit minority ethnic groups hardest, we would particularly like to hear the views of health and social care workers from minority ethnic backgrounds. 
 **ABOUT THIS SURVEY**
This research has received ethical approval from the London School of Hygiene & Tropical Medicine Ethics committee (Ref: 22923). The survey will take around 10 minutes to complete. Your participation is completely voluntary and you do not need to answer all the questions if you do not wish to. The survey is anonymous; we will not ask for any information in the survey that could directly identify who you are. All data will be stored securely at the London School of Hygiene & Tropical Medicine. 
 
Data from this survey will be made available in a public data repository for use by other researchers upon request. To access the data, researchers need to complete and submit a request form for the study investigators to consider. No data will be collected or shared which will allow for the direct identification of participants.
 
**AFTER THE SURVEY** 
At the end of the survey you will be given the opportunity to further voice your views regarding COVID-19 vaccination through a telephone or Zoom/Skype interview with one of the study investigators. If you decide you would like to take part in an interview, you will be asked to provide your contact details for us to get in touch with you. In this case, your contact details will not be linked to any of your survey answers.
  
**For more information about this research please feel free to contact me (Sadie Bell) and I will be happy to answer your questions. You can reach me on:****. These contact details are also provided at the end of the survey.**
 
 **Just here out of curiosity?**

Q2 **To take part in this survey, please confirm that you**

|  | **Yes** (1) | **No** (2) |
| --- | --- | --- |
| **1. Work in health or social care in the UK** (1) |  |  |
| **2. Have read and understood the study information above and consent to participate in this study** (2) |  |  |

Skip To: Q3 If To take part in this survey, please confirm that you [ No ] (Count) >= 1

Skip To: End of Block If To take part in this survey, please confirm that you [ Yes ] (Count) = 2

Q3
**Unfortunately, due to the inclusion criteria you are not eligible to take part in this survey**

 To take part in this survey you need to confirm that you:      1. Work in health or social care in the UK 
 2. Have read and understood the information on the first page of the survey and consent to participate in this study
 
If these criteria do apply to you then there is a chance that you selected the wrong option. If that is the case feel free to click the link and start again.     
 
**If you are not eligible to take part in this survey do you know someone that is?**
 
If you know someone who may be eligible to take part, please send them the link to the survey. This will really help us make the results of this survey as meaningful as possible. Thank you!   
 
**Study link:** <https://lshtm.qualtrics.com/jfe/form/SV_0pst28dLEW7z6dv>
 
If you have any questions about this project please feel free to contact the lead investigators - Sadie Bell on, Pauline Paterson on, or Sandra Mounier-Jack on.

Skip To: End of Survey If Unfortunately, due to the inclusion criteria you are not eligible to take part in this surveyTo t... Is Displayed

End of Block: Block 1

Start of Block: Block 2

Q4 **How would you describe your gender?**

- **Male** (1)
- **Female** (2)
- **Prefer to self describe (please enter the term you use to describe your gender)** (3) ________________________________________________
- **Prefer not to say** (4)

Q5 **What is your age?**

- **Under 25 years** (1)
- **25 to 34 years** (2)
- **35 to 44 years** (3)
- **45 to 54 years** (4)
- **55 to 64 years** (5)
- **65 years and over** (6)
- **Prefer not to say** (7)

Q6 **What religion, if any, do you most closely identify with?**

- **No religion** (1)
- **Atheist** (2)
- **Buddhist** (3)
- **Christian (including Church of England, Catholic, Protestant and all other Christian denominations)** (4)
- **Hindu** (5)
- **Jewish** (6)
- **Muslim** (7)
- **Sikh** (8)
- **Any other religion (please write here)** (9) ________________________________________________
- **Prefer not to say** (10)

07
**Are your day-to-day activities limited because of a health problem or disability which has lasted, or is expected to last, at least 12 months (including any problems related to old age)?**

- **Yes, limited a little** (1)
- **Yes, limited a lot** (2)
- **No** (3)
- **Prefer not to say** (4)

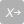

Q8 **What is your country of birth?**

▼ United Kingdom of Great Britain and Northern Ireland (185) ... Zimbabwe (1357)

9 **In the UK, people come from many different countries and cultures, and there are many different words to describe the different backgrounds or ethnic groups that people come from. The following questions are about your ethnicity or your ethnic group and how you feel about or identify with it.**      **Which of the following best describes your ethnicity?**

- **White:- British** (1)
- **White:- Irish** (2)
- **White:- Other White background** (4)
- **Gypsy or Irish Traveller** (3)
- **Mixed:- White and Black Caribbean** (5)
- **Mixed:- White and Black African** (6)
- **Mixed:- White and Asian** (7)
- **Mixed:- Other mixed background** (8)
- **Black or Black British:- African** (9)
- **Black or Black British:- Caribbean** (10)
- **Black or Black British:- Any other Black background** (11)
- **Asian or Asian British:- Indian** (12)
- **Asian or Asian British:- Pakistani** (13)
- **Asian or Asian British:- Bangladeshi** (14)
- **Asian or Asian British:- Other Asian background** (15)
- **Chinese** (16)
- **Arab** (17)
- **Other ethnic group not represented by these options (please state)** (18) ________________________________________________
- **Prefer not to say** (19)

10
**Please select how much you agree or disagree with the following statements.**

|  | **Strongly disagree** (1) | **Disagree** (2) | **Neither agree nor disagree** (3) | **Agree** (4) | **Strongly Agree** (5) |
| --- | --- | --- | --- | --- | --- |
| **I have spent time trying to find out more about my ethnic group, such as its history, traditions and customs** (1) |  |  |  |  |  |
| **I have a strong sense of belonging to my own ethnic group** (2) |  |  |  |  |  |
| **I have often done things that will help me to understand my ethnic group better** (3) |  |  |  |  |  |
| **I have often talked to other people to learn more about my ethnic group** (4) |  |  |  |  |  |
| **I feel a strong attachment towards my ethnic group** (5) |  |  |  |  |  |

End of Block: Block 2

Start of Block: Block 3

Q11 **Which best describes the sector that you most often work in?**

- **Health care** (1)
- **Social care** (2)

Q12 **How long have your worked in this sector?**

- **Less than 1 year** (1)
- **1-3 years** (2)
- **4-6 years** (3)
- **7-9 years** (4)
- **10-12 years** (5)
- **13-15 years** (6)
- **16 years or more** (7)

Q64 **In which country do you currently work?**

- **England** (1)
- **Scotland** (2)
- **Wales** (3)
- **Northern Ireland** (4)

Display This Question:

If In which country do you currently work? = <strong>England</strong>

Q13 **In which region of England do you currently work?**

- **South East** (1)
- **Greater London** (2)
- **North West** (3)
- **East of England** (4)
- **West Midlands** (5)
- **South West** (6)
- **Yorkshire and the Humber** (7)
- **East Midlands** (8)
- **North East** (9)
- **Prefer not to answer** (10)

Display This Question:

If Which best describes the sector that you most often work in? = <strong>Health care</strong>

Q14 **Which best describes the sector that you work in?**

- **Primary - Community** (1)
- **Primary - General practice** (2)
- **Secondary (hospital or community care that is planned (elective), or urgent and emergency) (3)**
- **Tertiary (highly specialised services - for people needing complex treatments who are referred from primary or secondary care) (4)**

Display This Question:

If Which best describes the sector that you most often work in? = <strong>Social care</strong>

Q15 **Please select your employer type**

- **Independent** (1)
- **Local authority** (2)
- **Jobs for direct payment recipients** (3)
- **NHS** (4)
- **Other (please write here)** (5) ________________________________________________

Display This Question:

If Which best describes the sector that you most often work in? = <strong>Social care</strong>

Q16 **Please select your role by main type of service group**

- **Residential** (1)
- **Domiciliary**  (2)
- **Day** (3)
- **Community** (4)
- **Other (please write here)** (5) ________________________________________________
- **Do not know** (6)

Display This Question:

If Which best describes the sector that you most often work in? = <strong>Social care</strong>

Q17 **What is your job role?**

- **Care worker** (1)
- **Senior care worker** (2)
- **Registered manager** (3)
- **Social worker (LA)**	(4)
- **Occupational therapist (LA)** (5)
- **Social care support staff** (6)
- **Registered nurse in social care** (7)
- **Other (please write here)** (8) ________________________________________________

Display This Question:

If Which best describes the sector that you most often work in? = <strong>Health care</strong>

Q18 **Which of the following best describes your occupational group?**

- **Allied Health Professional/Healthcare Scientist/Scientific and Technical** (1)
- **Medical and Dental** (2)
- **Ambulance (operational)** (3)
- **Public Health** (4)
- **Commissioning** (5)
- **Registered Nurses and Midwives** (6)
- **Nursing or Healthcare Assistants** (7)
- **Wider Healthcare Team** (8)
- **General Management** (9)

Display This Question:

If Which of the following best describes your occupational group? = <strong>Allied Health Professional/Healthcare Scientist/Scientific and Technical</strong>

Q19 **Which role do you currently work in?**

- **Occupational Therapy** (1)
- **Physiotherapy** (2)
- **Radiography** (3)
- **Pharmacy** (4)
- **Clinical Psychology** (5)
- **Psychotherapy** (6)
- **Operating Department Practitioner** (7)
- **Other qualified Allied Health Professionals (e.g. dietetics, speech and language therapy)** (8)
- **Support to Allied Health Professionals (e.g. support worker, therapy helper, therapy assistant or student)** (9)
- **Other qualified Scientific and Technical or Healthcare Scientists (e.g. haematology, clinical biochemistry, microbiology)** (10)
- **Support to Healthcare Scientists (e.g. technicians, assistants or students)** (11)
- **Other (please write here)** (12) ________________________________________________

Display This Question:

If Which of the following best describes your occupational group? = <strong>Medical and Dental</strong>

Q20 **Which role do you currently work in?**

- **Medical - Consultant** (1)
- **Dental - Consultant** (2)
- **Medical - In Training**	(3)
- **Dental - In Training** (4)
- **Salaried Primary Care Dentists** (5)
- **Other (please write here)** (6) ________________________________________________
- **Medical - General practitioner** (7)
- **Medical - Other (e.g. Staff and Associate Specialists / Non-consultant career grade)** (8)
- **Dental - Other (e.g. Staff and Associate Specialists / Non-consultant career grade)** (9)

Display This Question:

If Which of the following best describes your occupational group? = <strong>Ambulance (operational)</strong>

Q21 **Which role do you currently work in?**

- **Emergency Care Practitioner** (1)
- **Paramedic** (2)
- **Emergency Care Assistant**	(3)
- **Ambulance Technician** (4)
- **Ambulance Control Staff (e.g. call handler, dispatchers, PTS controllers)** (5)
- **Patient Transport Service (e.g. ambulance drivers, support staff)** (6)
- **Other (please write here)** (7) ________________________________________________

Display This Question:

If Which of the following best describes your occupational group? = <strong>Registered Nurses and Midwives</strong>

Q22 **Which role do you currently work in?**

- **Nurse - Adult/General** (1)
- **Nurse - Mental health** (2)
- **Nurse - Learning disabilities** (3)
- **Nurse - Children** (4)
- **Nurse - District/Community** (5)
- **Nurse - Other (please write here)** (6) ________________________________________________
- **Midwife** (7)
- **Health Visitor** (8)
- **Other (please write here)** (9) ________________________________________________

Display This Question:

If Which of the following best describes your occupational group? = <strong>Wider Healthcare Team</strong>

Q23 **Which role do you currently work in?**

- **Admin & Clerical (including Medical Secretary)** (1)
- **Central Functions/Corporate Services (e.g. HR, Finance, Information Systems, Information Technology)**	(2)
- **Maintenance/Ancillary (e.g. housekeeping, domestic staff, maintenance, facilities, estates)** (3)
- **Other (please write here)** (4) ________________________________________________

Display This Question:

If Which best describes the sector that you most often work in? = <strong>Health care</strong>

Q24 **What pay band (Agenda for Change) are you currently working at?**

- **Band 1** (1)
- **Band 2** (2)
- **Band 3** (3)
- **Band 4** (4)
- **Band 5** (5)
- **Band 6** (6)
- **Band 7** (7)
- **Band 8a** (8)
- **Band 8b** (9)
- **Band 8c** (10)
- **Band 8d** (11)
- **Band 9** (12)
- **Not applicable - not within the Agenda for Change pay system** (13)
- **Do not know**	(14)

Q25 **To what extent do you agree with the following:**

**I would recommend my organisation as a place to work**

- **Strongly agree** (1)
- **Somewhat agree** (2)
- **Neither agree nor disagree** (3)
- **Somewhat disagree** (4)
- **Strongly disagree** (5)

Q55 **In the last 6 months have you personally experienced discrimination (being treated less favourably by someone else because of your ethnic background, gender, religion, sexual orientation, disability, age) at work from any of the following?**

|  | **Yes** (1) | **No** (2) |
| --- | --- | --- |
| **Patients, service users, or clients** (1) |  |  |
| **Colleagues** (2) |  |  |

Display This Question:

If In the last 6 months have you personally experienced discrimination (being treated less favourabl... [ Yes ] (Count) >= 1

Q56 **If you have experienced any discrimination at work, on what grounds have you experienced discrimination?**

- **Ethnic background** (1)
- **Gender** (2)
- **Religion** (3)
- **Sexual orientation** (4)
- **Disability** (5)
- **Age** (6)
- **Other** (7)

| Page Break |
| --- |

Q26 **On an average day in your job, how much of your time is spent in direct (i.e. face-to-face) contact with patients, service users, or clients?**

- **Almost all** (1)
- **Around half** (2)
- **Less than half** (3)
- **None** (4)

Q27 **In your job role, do you come into direct (i.e. face-to-face) contact with patients, service users, or clients at higher risk of severe COVID-19? (e.g. due to older age or clinical risk)**

- **Yes** (1)
- **No** (2)
- **Unsure** (3)

Q52 **In your job role, to your knowledge have you provided direct care (i.e. had face-to-face contact) with a patient, service user, or client diagnosed with COVID-19?**

- **Yes** (1)
- **No** (2)

Q28 **Do you belong to a group that is considered at higher risk of severe COVID-19?**

- **Yes** (1)
- **No** (2)
- **Unsure** (3)

Q29
**Have you ever tested positive for COVID-19 during the pandemic?**

- **Yes – I have tested positive for COVID-19** (1)
- **No – I have either not been tested, or my test(s) was negative or inconclusive** (2)

Display This Question:

If Have you ever tested positive for COVID-19 during the pandemic? = <strong>No – I have either not been tested, or my test(s) was negative or inconclusive</strong>

Q63 **Although you have never tested positive for COVID-19, do you think you may have had COVID-19 at some stage during the pandemic?**

- **Yes** (1)
- **No** (2)

Q62 **To your knowledge, has a close friend or family member of yours tested positive for COVID-19 at any point during the pandemic?**

- **Yes** (1)
- **No** (2)

Q31 **Have you had to self-isolate during the pandemic due to potential COVID-19 exposure/displaying potential COVID-19 symptoms?**

- **Yes** (1)
- **No** (2)

Display This Question:

If Have you had to self-isolate during the pandemic due to potential COVID-19 exposure/displaying po... = <strong>Yes</strong>

Q71 **How many times have you had to self-isolate during the pandemic?**

- **Once** (1)
- **Twice** (2)
- **3 times** (3)
- **4 or more times** (4)

| Page Break |
| --- |

Q32 **How much has the COVID-19 pandemic impacted on your day-to-day life?**

- **No impact** (1)
- **Minor impact** (2)
- **Moderate impact** (3)
- **Major impact** (4)

End of Block: Block 3

Start of Block: Block 3

Q35 **This section of the survey will focus on vaccinations for health and social care workers**
 
**Have you had the seasonal flu vaccine for this season (2020/21)?**

- **Yes** (1)
- **No** (2)

Q68 **Did you had the seasonal flu vaccine for the previous season (2019/20)?**

- **Yes** (1)
- **No** (2)

| Page Break |
| --- |

Display This Question:

If Which best describes the sector that you most often work in? = <strong>Social care</strong>

Q65 **Please indicate your level of agreement with each of the following statements**

|  | **Strongly agree** (1) | **Tend to agree** (2) | **Tend to disagree** (3) | **Strongly disagree** (4) | **Don't know** (5) |
| --- | --- | --- | --- | --- | --- |
| **I think COVID-19 is deadlier than seasonal flu** (1) |  |  |  |  |  |
| **I think it's important for social care workers to get a COVID-19 vaccine to protect themselves** (2) |  |  |  |  |  |
| **I think it’s important for social care workers to get a COVID-19 vaccine to protect their families** (3) |  |  |  |  |  |

Display This Question:

If Which best describes the sector that you most often work in? = <strong>Social care</strong>

Q63 **Please indicate your level of agreement with each of the following statements**

|  | **Strongly agree** (1) | **Tend to agree** (2) | **Tend to disagree** (3) | **Strongly disagree** (4) | **Don't know** (5) |
| --- | --- | --- | --- | --- | --- |
| **I think it’s important for social care workers to get a COVID-19 vaccine to protect their patients, service users, or clients** (4) |  |  |  |  |  |
| **I think that COVID-19 vaccines are safe** (5) |  |  |  |  |  |
| **I think that COVID-19 vaccines are effective** (6) |  |  |  |  |  |

Display This Question:

If Which best describes the sector that you most often work in? = <strong>Social care</strong>

Q64 **Please indicate your level of agreement with each of the following statements**

|  | **Strongly agree** (1) | **Tend to agree** (2) | **Tend to disagree** (3) | **Strongly disagree** (4) | **Don't know** (5) |
| --- | --- | --- | --- | --- | --- |
| **I think it is important for people to get vaccinated against COVID-19 to get life back to 'normal'** (7) |  |  |  |  |  |
| What religion, if any, do you most closely identify with? != <strong>No religion</strong>  **I think that COVID-19 vaccines are compatible with my religious beliefs** (8) |  |  |  |  |  |
| **I feel well informed about COVID-19 vaccination** (9) |  |  |  |  |  |

Display This Question:

If Which best describes the sector that you most often work in? = <strong>Health care</strong>

Q66 **Please indicate your level of agreement with each of the following statements**

|  | **Strongly agree** (1) | **Tend to agree** (2) | **Tend to disagree** (3) | **Strongly disagree** (4) | **Don't know** (5) |
| --- | --- | --- | --- | --- | --- |
| **I think COVID-19 is deadlier than seasonal flu** (1) |  |  |  |  |  |
| **I think it's important for health care workers to get a COVID-19 vaccine to protect themselves** (2) |  |  |  |  |  |
| **I think it’s important for health care workers to get a COVID-19 vaccine to protect their families** (3) |  |  |  |  |  |

Display This Question:

If Which best describes the sector that you most often work in? = <strong>Health care</strong>

Q67 **Please indicate your level of agreement with each of the following statements**

|  | **Strongly agree** (1) | **Tend to agree** (2) | **Tend to disagree** (3) | **Strongly disagree** (4) | **Don't know** (5) |
| --- | --- | --- | --- | --- | --- |
| **I think it’s important for health care workers to get a COVID-19 vaccine to protect their patients, service users or clients** (4) |  |  |  |  |  |
| **I think that COVID-19 vaccines are safe** (5) |  |  |  |  |  |
| **I think that COVID-19 vaccines are effective** (6) |  |  |  |  |  |

Display This Question:

If Which best describes the sector that you most often work in? = <strong>Health care</strong>

Q68 **Please indicate your level of agreement with each of the following statements**

|  | **Strongly agree** (1) | **Tend to agree** (2) | **Tend to disagree** (3) | **Strongly disagree** (4) | **Don't know** (5) |
| --- | --- | --- | --- | --- | --- |
| **I think it is important for people to get vaccinated against COVID-19 to get life back to 'normal'** (7) |  |  |  |  |  |
| What religion, if any, do you most closely identify with? != <strong>No religion</strong>  **I think that COVID-19 vaccines are compatible with my religious beliefs** (8) |  |  |  |  |  |
| **I feel well informed about COVID-19 vaccination** (9) |  |  |  |  |  |

Q36 **Now that a COVID-19 vaccine has become available to health and social care workers, have you been offered a COVID-19 vaccine?**

- **Yes** (1)
- **No** (2)

Display This Question:

If Now that a COVID-19 vaccine has become available to health and social care workers, have you been... = <strong>Yes</strong>

Q37 **Have you had or are you intending to have the COVID-19 vaccine?**

- **Yes, I have had the vaccine** (1)
- **I have been offered the vaccine and have or will book a vaccination appointment** (2)
- **I have been offered the vaccine but have decided not to take it** (3)

Display This Question:

If Now that a COVID-19 vaccine has become available to health and social care workers, have you been... = <strong>Yes</strong>

Q65 **Which COVID-19 vaccine were you offered?**

- **Pfizer/BioNTech vaccine** (1)
- **Oxford/AstraZeneca vaccine** (2)
- **Moderna vaccine** (3)
- **Other (please state)** (5) ________________________________________________
- **Do not know** (4)

Display This Question:

If Have you had or are you intending to have the COVID-19 vaccine?  = <strong>Yes, I have had the vaccine</strong>

Or Have you had or are you intending to have the COVID-19 vaccine?  = <strong>I have been offered the vaccine and have or will book a vaccination appointment</strong>

Q38 **Please can you tell us the main reason you decided to get the vaccine?**

________________________________________________________________

________________________________________________________________

________________________________________________________________

________________________________________________________________

________________________________________________________________

Display This Question:

If Have you had or are you intending to have the COVID-19 vaccine?  = <strong>I have been offered the vaccine but have decided not to take it</strong>

Q70 **Please can you tell us the main reason you decided not to get the vaccine?**

________________________________________________________________

________________________________________________________________

________________________________________________________________

________________________________________________________________

________________________________________________________________

Display This Question:

If Have you had or are you intending to have the COVID-19 vaccine?  = <strong>Yes, I have had the vaccine</strong>

Q53 **How easy or difficult was it to access the COVID-19 vaccine in your organisation?**

|  | 1 (1) | 2 (2) | 3 (3) | 4 (4) | 5 (5) | 6 (6) |  |
| --- | --- | --- | --- | --- | --- | --- | --- |
| **Very easy** |  |  |  |  |  |  | **Very difficult** |

Display This Question:

If Now that a COVID-19 vaccine has become available to health and social care workers, have you been... = <strong>No</strong>

Q39
**If you were offered a COVID-19 vaccine would you accept it?**

- **Yes, definitely**	(1)
- **Unsure but leaning towards yes**	(2)
- **Unsure but leaning towards no** (3)
- **No, definitely not** (4)

Display This Question:

If If you were offered a COVID-19 vaccine would you accept it?  = <strong>Yes, definitely </strong>

Or If you were offered a COVID-19 vaccine would you accept it?  = <strong>Unsure but leaning towards yes </strong>

Or If you were offered a COVID-19 vaccine would you accept it?  = <strong>Unsure but leaning towards no</strong>

Or If you were offered a COVID-19 vaccine would you accept it?  = <strong>No, definitely not</strong>

Q40 **Please can you tell us the main reason why?**

________________________________________________________________

________________________________________________________________

________________________________________________________________

________________________________________________________________

________________________________________________________________

| Page Break |
| --- |

Display This Question:

If Have you had or are you intending to have the COVID-19 vaccine?  = <strong>I have been offered the vaccine but have decided not to take it</strong>

Or Have you had or are you intending to have the COVID-19 vaccine?  = <strong>I have been offered the vaccine and have or will book a vaccination appointment</strong>

Or Now that a COVID-19 vaccine has become available to health and social care workers, have you been... = <strong>No</strong>

Q60 **Please indicate your level of agreement with each of the following statements**

|  | **Strongly agree** (1) | **Tend to agree** (2) | **Somewhat disagree** (3) | **Strongly disagree** (4) | **Don't know** (5) |
| --- | --- | --- | --- | --- | --- |
| **My family and friends expect me to accept a COVID-19 vaccine** (1) |  |  |  |  |  |
| **My colleagues expect me to accept a COVID-19 vaccine** (2) |  |  |  |  |  |
| **I feel under pressure from my employer to get a COVID-19 vaccine** (3) |  |  |  |  |  |
| **I am worried about getting side-effects from a COVID-19 vaccine** (4) |  |  |  |  |  |

Display This Question:

If Have you had or are you intending to have the COVID-19 vaccine?  = <strong>I have been offered the vaccine and have or will book a vaccination appointment</strong>

Or Have you had or are you intending to have the COVID-19 vaccine?  = <strong>I have been offered the vaccine but have decided not to take it</strong>

Or Now that a COVID-19 vaccine has become available to health and social care workers, have you been... = <strong>No</strong>

Q54 **How easy or difficult do you think it is to access the COVID-19 vaccine in your organisation?**

|  | 1 (1) | 2 (2) | 3 (3) | 4 (4) | 5 (5) | 6 (6) |  |
| --- | --- | --- | --- | --- | --- | --- | --- |
| **Very easy** |  |  |  |  |  |  | **Very difficult** |

Display This Question:

If Have you had or are you intending to have the COVID-19 vaccine?  = <strong>Yes, I have had the vaccine</strong>

Q61 **Please indicate your level of agreement with each of the following statements**

|  | **Strongly agree** (1) | **Tend to agree** (2) | **Tend to disagree** (3) | **Strongly disagree** (4) | **Don't know** (5) |
| --- | --- | --- | --- | --- | --- |
| **My family and friends expected me to accept a COVID-19 vaccine** (1) |  |  |  |  |  |
| **My colleagues expected me to accept a COVID-19 vaccine** (2) |  |  |  |  |  |
| **I felt under pressure from my employer to get a COVID-19 vaccine** (3) |  |  |  |  |  |
| **I was worried about getting side-effects from a COVID-19 vaccine** (4) |  |  |  |  |  |

| Page Break |
| --- |

Display This Question:

If Now that a COVID-19 vaccine has become available to health and social care workers, have you been... = <strong>No</strong>

Or Have you had or are you intending to have the COVID-19 vaccine?  = <strong>I have been offered the vaccine and have or will book a vaccination appointment</strong>

Or Have you had or are you intending to have the COVID-19 vaccine?  = <strong>I have been offered the vaccine but have decided not to take it</strong>

Q58 **How much would you worry about contracting COVID-19 in the next 6 months if you were not to get a COVID-19 vaccine?**

- **A great deal** (1)
- **A lot** (2)
- **A moderate amount** (3)
- **A little** (4)
- **None at all** (5)

Display This Question:

If Now that a COVID-19 vaccine has become available to health and social care workers, have you been... = <strong>No</strong>

Or Have you had or are you intending to have the COVID-19 vaccine?  = <strong>I have been offered the vaccine and have or will book a vaccination appointment</strong>

Or Have you had or are you intending to have the COVID-19 vaccine?  = <strong>I have been offered the vaccine but have decided not to take it</strong>

Q57 **How much regret would you feel in the next 6 months if you did not get a COVID-19 vaccine and subsequently contracted COVID-19?**

- **A great deal** (1)
- **A lot** (2)
- **A moderate amount** (3)
- **A little** (4)
- **None at all** (5)

| Page Break |
| --- |

Q70 **We are interested in which sources of information you trust. Please can you tell us the level of agreement you have with the following statements.**
 
**I trust the advice on Covid-19 vaccination given by...**

|  | **Strongly agree** (1) | **Agree** (2) | **No opinion** (3) | **Disagree** (4) | **Strongly disagree** (5) |
| --- | --- | --- | --- | --- | --- |
| **My work colleagues** (1) |  |  |  |  |  |
| **Social media** (2) |  |  |  |  |  |
| **Community leaders** (3) |  |  |  |  |  |
| **Religious leaders** (4) |  |  |  |  |  |
| **NHS** (5) |  |  |  |  |  |
| **News media (e.g. print or online newspapers, radio and television news broadcasts)** (6) |  |  |  |  |  |
| **Government** (7) |  |  |  |  |  |
| **Family** (8) |  |  |  |  |  |
| **Friends** (9) |  |  |  |  |  |
| **Scientists involved in COVID-19 vaccine development** (10) |  |  |  |  |  |
| **Public Health England** (11) |  |  |  |  |  |
| **Health professionals** (12) |  |  |  |  |  |

| Page Break |
| --- |

Q46 **Do you have anything further you would like to add about your views on COVID-19 vaccination for health or social care workers? (If yes, please use this text box)**

________________________________________________________________

________________________________________________________________

________________________________________________________________

________________________________________________________________

________________________________________________________________

End of Block: Block 3

Start of Block: Block 2

Q48 Thank you for completing this survey! The study that you have just participated in looks at health and social care workers views on COVID-19 vaccination. The results of this study will be communicated to Public Health England and may inform the communication and delivery of the COVID-19 vaccination programme to health and social care workers.
 **Follow up telephone or Zoom/Skype interview** For this study we are planning to combine the results of this survey with the findings from a range of interviews with health and social care workers. If you would like to help us further and would be willing to participate in an approximately 30-45 mins long conversation on a similar topic as that covered in this survey please provide, your email address or mobile phone details in the boxes below. Alternatively you can contact me directly on:. As a thank you for taking part in interviews, participants will receive a **£10 multi-store gift voucher.**

|  | (1) |
| --- | --- |
| **Email address** (1) |  |
| **Phone number** (2) |  |

Q49 **Additional details**     This study forms part of the research agenda of the National Institute for Health Research (NIHR) Health Protection Research Unit in Immunisation. Please contact Sadie Bell, Pauline Paterson, or Sandra Mounier-Jack for further information about the study.   Help us make the results of this project as meaningful as possible   Thank you for participating in this survey. If you have signed up for an interview, thank you and you will be hearing from one of our investigators shortly.   In the meantime, however there is a good chance that you currently know other frontline health and social care workers. If you know of someone who may be eligible to take part in this survey, and think that they would like to participate, please do pass the word on about this study by sending them the link highlighted below directly.   The more people that take this survey the more meaningful the results become so any help you can give would be very much appreciated! Thank you.   The link to this study is: <https://lshtm.qualtrics.com/jfe/form/SV_0pst28dLEW7z6dv>

End of Block: Block 2
