## Supplementary material for "COVID-19 vaccination beliefs, attitudes, and behaviours among health and social care workers in the UK: a mixed-methods study": S1 Appendix 2

### Supplemental material – recoding variables

#### 1. Combining categorical variables for analysis

##### 1.1. Ethnicity

The following outlines how each of the Ethnicity subcategories, used in the statistical analysis, were constructed from the original survey question. **(1)** **White British and White Irish** is combination of White British (n=1051) and White Irish (n=51). **(2) White Other** is a combination of Gypsy or Irish Traveller(n=1), White Other White Backgrounds (n=93). **(3)** **Black or Black British African or Mixed Black African** is a combination of Black and Black British African (n=155) and Mixed White and Black African (n=13). **(4) Black or Black British Caribbean or Mixed Black Caribbean** is a combination of Black or Black British Caribbean (n=51) and Mixed White and Black Caribbean (n=15). **(5) Asian or Asian British Indian** was a single answer in the original questions (n=264). **(6) Other South East Asian or Mixed Asian** is a combination of Asian or Asian British Pakistani (n=32), Asian or Asian British Bangladeshi (n=11), Asian or Asian British Other Asian Backgrounds (n=50), and Mixed White and Asian (n=16). **(7) Other Ethnic Minorities** is a combination of Arab (n=8), Chinese (n=14), Black or Black British any other Black Background (n=15), Mixed other mixed background (n=19) and Other ethnic minority not represented by these options (n=34).

##### 1.2. Job role

The following outlines how each of the Job role subcategories, used in the statistical analysis, were constructed from the original survey question. **(1) Allied Health Professionals** was a single answer in the original questions **(2) Medical** was a single answer in the original questions **(3) Registered Nursing and Midwives** was a single answer in the original questions **(4) Nursing or Healthcare** assistant was a single answer in the original questions **(5) Social Care** all participants that indicated that their sector was social care as there was not enough data to delimitate further **(6) Other** is a combination of Ambulance (operational), Public Health, Commissioning, Wider Healthcare Team and General Management.

#### 2. Combining continuous variables

##### 2.1. COVID-19 vaccine attitudes and beliefs

To reduce the number of attitude and belief variables included in the COVID-19 vaccine uptake regression analysis we conducted a factor analysis to determine which items were suitable to combine into a single variable. Below and Table 1 outline the results of a factor analysis that we conducted on these variables.

Component 1: Eigenvalue = 5.453, Variance = 45.4

Component 2: Eigenvalue = 1.308, Variance = 10.9

Component 3: Eigenvalue = 1.201, Variance = 10.0

##### Table 1: Pattern Matrix for the three components identified in factor analysis (principle components analysis)

| **Statement** | **Component 1** | **Component 2** | **Component 3** |
| --- | --- | --- | --- |
| *“I think COVID-19 is deadlier than seasonal flu”* | 0.534 | 0.377 | 0.170 |
| *“I think it's important for social/health care workers to get a COVID-19 vaccine to protect themselves”* | 0.878 | 0.312 | -0.001 |
| *“I think it’s important for social/health care workers to get a COVID-19 vaccine to protect their families”* | 0.859 | 0.365 | -0.007 |
| *“I think it’s important for social/health care workers to get a COVID-19 vaccine to protect their patients”* | 0.838 | 0.329 | 0.020 |
| *“I think that COVID-19 vaccines are safe”* | 0.802 | -0.207 | -0.139 |
| *“I think that COVID-19 vaccines are effective”* | 0.764 | -0.175 | -0.116 |
| *“I think it is important for people to get vaccinated against COVID-19 to get life back to 'normal'”* | 0.773 | 0.137 | -0.017 |
| *“I feel well informed about COVID-19 vaccination”* | 0.597 | -0.385 | -0.135 |
| *“My family and friends expect me to accept a COVID-19 vaccine”* | 0.559 | -0.330 | 0.490 |
| *“My colleagues expect me to accept a COVID-19 vaccine”* | 0.372 | -0.406 | 0.694 |
| *“I felt under pressure from my employer to get a COVID-19 vaccine”* | -0.352 | 0.195 | 0.539 |
| *“I am worried about getting side-effects from a COVID-19 vaccine”* | -0.450 | 0.524 | 0.328 |

After discussion with co-authors it was decided that the following items could be combined into a single variable: **(1)** *“I think it's important for social/health care workers to get a COVID-19 vaccine to protect themselves”* **(2)** *“I think it’s important for social/health care workers to get a COVID-19 vaccine to protect their families”* **(3)** *“I think it’s important for social/health care workers to get a COVID-19 vaccine to protect their patients”* **(4)** *“I think that COVID-19 vaccines are safe”* **(5)** *“I think that COVID-19 vaccines are effective”* **(6)** *“I think it is important for people to get vaccinated against COVID-19 to get life back to 'normal'”.* This new variable was assigned the label: **Combined COVID-19 vaccine beliefs (important, safe, and effective)**

Similarly, the items *“My family and friends expect me to accept a COVID-19 vaccine”* and *“My colleagues expect me to accept a COVID-19 vaccine”* were combined to form a variable we labelled **Social norms to vaccinate against COVID-19.** Initial we included *“I felt under pressure from my employer to get a COVID-19 vaccine”* with in this combined variable but later separated it and used it as a single item variable due to the importance participants placed on pressure within qualitative interviews.

*“I think COVID-19 is deadlier than seasonal flu”*, *“I feel well informed about COVID-19 vaccination”*, and *“I am worried about getting side-effects from a COVID-19 vaccine”* were kept as single item variables in the analysis due to them not clearly loading on to any of the components suggested by the factor analysis.

##### 2.2. Trust in sources of information

To reduce the number of trust variables included in the COVID-19 vaccine uptake regression analysis we conducted a factor analysis to determine which items were suitable to combine into a single variable. Below and Table 2 outline the results of a factor analysis that we conducted on these variables.

Component 1: Eigenvalue = 4.768, Variance = 39.7

Component 2: Eigenvalue = 2.07, Variance = 17.2

Component 3: Eigenvalue = 1.033, Variance = 8.6

Table 2: Pattern Matrix for the three components identified in factor analysis (principle components analysis)

| **I trust the advice on Covid-19 vaccination given by…** | **Component 1** | **Component 2** | **Component 3** |
| --- | --- | --- | --- |
| *My work colleagues* | 0.313 | 0.060 | -0.482 |
| *Social media* | -0.218 | 0.734 | -0.111 |
| *Community leaders* | 0.149 | 0.679 | -0.120 |
| *Religious leaders* | -0.072 | 0.731 | -0.129 |
| *NHS* | 0.842 | 0.078 | -0.004 |
| *News media (e.g. print or online newspapers, radio, and television news broadcasts)* | 0.243 | 0.636 | 0.030 |
| *Government* | 0.571 | 0.438 | 0.168 |
| *Family* | -0.012 | 0.130 | -0.833 |
| *Friends* | -0.023 | 0.080 | -0.889 |
| *Scientists involved in COVID-19 vaccine development* | 0.828 | -0.117 | -0.096 |
| *Public Health England* | 0.852 | 0.072 | 0.088 |
| *Health Professionals* | 0.816 | -0.124 | -0.233 |

After decision with co-authors the following combination of items were suggested for use in the COVID-19 vaccine uptake regression analysis.

**Trust in health system sources**. Containing: *NHS*, *Scientists involved in COVID-19 vaccine development*, *Public Health England*, and *Health Professionals*.

**Trust in non-health system sources**. Containing: *Social Media*, *Community leaders*, *Religious leaders,* and *News media.*

**Trust in Friends and Family members**. Containing: *Family* and *Friends*.

The following two items did not clearly load onto any of the three components suggested by the factor analysis: **(1)** *My work colleagues*, and **(2)** *Government*. As such we included these as single item variables within our analysis.
